# Tele- Yoga therapy for Patients with Chronic Pain during Covid-19 Lockdown: A Prospective Nonrandomized Single Arm Clinical Trial

**DOI:** 10.1101/2020.07.16.20154229

**Authors:** Neha Sharma, Dipa Modi, Asha Nathwani, Bhavna Pandya, Jaydeep Joshi

**Author notes:** Corresponding author: Dr. Neha Sharma; Aarogyam (UK) CIC, 143 Loughborough Road, Leicester, UK.

## Abstract

**Background:** Pain management services and support programs have been closed during pandemic. Self-management options, particularly for chronic pain, is required which can be undertaken at one’s own convenience and without leaving home.

**Objectives:** To evaluate the impact of tele-yoga therapy on patients suffering with chronic pain reducing pain intensity, disability, anxiety and depression.

**Material and methods:** In total 18 patients with different chronic pain diagnosis were recruited to individual yoga Therapy sessions twice a week at home (tele-yoga) using a videoconference app. Each participant followed set of practices every day at home. Main outcome measures included pain intensity, pain disability, anxiety and depression scores. Data were collected at baseline and after 6-weeks of intervention.

**Results:** There were significant improvement in pain intensity from Baseline to 6-weeks (P<0.001); also pain disability (P<0,001). Both scores of anxiety and depression reduced at the end of intervention period (P<0,001; P<0,001).

**Conclusions:** Pilot results suggest that tele-yoga therapy may be an effective tool to self-manage chronic pain and related functional and psychological impacts. Further larger studies, randomized, controlled trials are needed to confirm the preliminary outcome.

**Trial Registration:** ClinicalTrials.gov NCT04457388; https://clinicaltrials.gov/ct2/show/NCT04457388

## Introduction

Around the world, chronic pain is a cause for significant suffering, functional limitation, and poor quality of life^1,2^. Recent COVID-19 outbreak led to closing all elective procedures, outpatients’ services including chronic pain management services for limiting the spread of virus. Patients were also requested to strict isolation, stay at home and social distancing.^3^ Chronic pain conditions are often associated with impaired quality of life due to significant physical and psychological burden. With major psychological impact and isolation imposed by the pandemic may worsen pain, mental health and overall quality of life. This may in turn significantly rise the chronic pain and co-morbid conditions.^4^

Previous studies demonstrated that yoga could improve pain measures^5^, and physical function^6^ as well as reduce symptoms of anxiety and depression in people with chronic pain^7,8^. Yoga Therapy is a form of Yoga to treat medical condition. For chronic pain, multidimensional approach of Yoga therapy is used, from the physical, emotional, mental and spiritual to highest self-awareness^9^. It includes all methods of physical postures, breath control, sensory methods, affirmations and visualisations, chants and meditation. In traditions, yoga therapy is primarily a science of self-healing aimed at relieving the disease of body and mind.^10^

Tele-health or telemedicine uses telecommunications technologies to have increased access to health care^11^. Telemedicine have been used to provide support for chronic pain patients during pandemic^12^. Previous studies have found telehealth interventions for chronic pain feasible and possibly effective, including pain management,^13^ physical exercise,^14^ and psychological interventions.^15,16^ Tele-yoga intervention has only been used in one study having patients with heart failure and chronic obstructive pulmonary disease,^17^ which found intervention acceptable and applicable in other chronically ill patients.

To date, there has been no previous study using tele-yoga therapy in chronic pain management. Therefore, present study was designed to test yoga therapy sessions conveyed through a technical solution remotely on chronic pain patients.

## Methods

Present study was a single arm nonrandomized clinical trial to investigate the impact of tele-yoga therapy on patients with chronic pain. We specifically examined the efficacy of tele-yoga therapy for pain relief, pain related disability, anxiety, and depression.

Patients were recruited from Leicester, UK through flyers, social media, referrals from local surgery and complementary and alternative medicine practitioners. Patients were included in the study if they were: 18 years of age to 60 years, having chronic nonmalignant pain for at least 3 months, having access to internet and video calls. Patients were excluded if pregnant or breast-feeding, not willing to give written consent and patients with severe psychiatric or personality disorder.

### Study design

Prospective study with 6-weeks interventions period was conducted by Aarogyam, (UK) in Leicester, UK between April 2020 and June 2020. Convenience sample of chronic pain patients were considered eligible and were enrolled in the study after their written informed consent. Patients were allowed to withdraw from the study at any point.

WhatsApp and zoom were found commonly used and easy to access by most participants. Therefore, WhatsApp and Zoom video calls were used according to each participants convenience, starting from completing baseline assessment at first session and then all the participants were booked for twice weekly tele-yoga therapy sessions.

All the patients provided written informed consent; ethics approval was taken from Independent Review Board. Procedures were in accordance with the ethical standards of the responsible committee on human experimentation and with the Helsinki Declaration of 2008.

### Intervention

Yoga therapy for individualized based on each participant personal needs and clinical conditions. Sessions were scheduled twice in a week with trained and experienced yoga therapist over the video conference for 45 minutes.

Each session included set of yoga practices combining yoga loosening exercises (sukshma Vyayama), Postures (Asana), Breathing exercises (Pranayama), Relaxation techniques and chanting (A, U, M sylabal chants) meditation. Each session, participants were given home practice sets to follow every day.

### Outcome measures

At baseline and post intervention, patients were given pain and practice diary to log daily practices, time to practice and pain score. Specific questionnaires to evaluate intensity of pain, ability to perform normal daily activities, depression and anxiety symptoms were taken before and after 6-week of intervention.

Pain intensity: visual-analogic scale (VAS), using pain level from 0 value (no pain) to 10 value (worst imaginable pain) evaluated pain intensity at baseline and end of 6-weeks.

Pain Disability Index (PDI): PDI used to measure the disability caused by chronic pain. It consists of seven rating scales, structured in Likert form, from 0 value (no disability) to 10 value (worst disability). Life activities are listed on each 7 categories and participants need to circle the number on the scale that described the level of disability they experience.

Hospital Anxiety and Depression Scale (HADS): The HADS is a self-assessment scale has been used to assess anxiety, depression and emotional distress among patients. It is composed by fourteen items, seven relating to anxiety and seven to depression. Each item scores from 0 to 3 values

Between 0-10 scale of program satisfaction, usefulness and technology rating were asked where 0 was worst and 10 was best.

### Statistical methods

MedCalc Statistical Software version 19.3.1 (MedCalc Software Ltd, Ostend, Belgium; https://www.medcalc.org; 2020) was used to conduct statistical analysis. It was the first pilot study, so power analysis was not considered. All the values are shown in numbers (%) and Mean ± Standard Deviation. A preliminary study of distribution with Shapiro-Wilk test showed that scores were normally distributed. So, Paired Students’ t test was used for variables. Graphics show mean values because we used parametric tests for the statistical analysis. Confidence interval is at 95%.

## Results

Twenty-two participants were recruited in initial contact. Of these, twenty participants met the eligibility criteria. One had to discontinue due to death in the family and one of the participants dropped out prior to the study end and was unavailable for post assessment. The 18 participants completed at least 10 of the 12 classes offered.

Our sample was composed of 66% women and 34% men with an average age of 47.28 ± 8.51 years old (21-58 years old), affected by non-specific chronic neck pain, back pain and knee pain, fibromyalgia, headache, arthritis characterized by chronic pain (Tab. 1). There was no significant difference reported at baseline in pain intensity, disability, anxiety and depression scales among patients with different medical condition.

Most of participants were Asian (77.7%), White (British) (11.1%) and Black (African) (11.2%). Out of Asian, most participants were Indian (61.1 %), Pakistani (11.1%) and Bangladeshi (5.5%). Four participants (22.2%) were self-referred from the social media, 9 (50%) were referred by someone who has used the pain services before and 5 (27.8%) were referred by local health services and practitioners.

### Intervention acceptability

All the participants reported positive experience of doing yoga at home in their own convenience. Home practices were given twice daily for 15-20 mins each time. All the participants found this helpful to have short time at once (9.6±0.98).

Yoga postures were modified as per each participant and their feasibility. Patients satisfaction was reported at the end was very much satisfied (9.2 ± 0.89). One participant (5.5%) had difficulty following some of the practices of deep relaxation over the video conference, so video instructions were sent to follow during home practice.

Eight participants (44.4%) used zoom for yoga session and 10 participants (55.6%) were comfortable using WhatsApp video chat. Zoom was better rated (8.2 ± 0.88) by participants than WhatsApp (7.2 ± 0.78) with interference, connection and visibility issues. However, app was decided upon participants choice and convenience.

### Outcome measure

Significant reduction in anxiety was reported in participants from baseline (11.72 ± 3.54) to post intervention (5.61± 2.03), along with depression (13.05 ± 3.22 vs 5.38 ± 1.33) (p<0.0001). (Table 2, Figure 1)

**TABLE 1:**
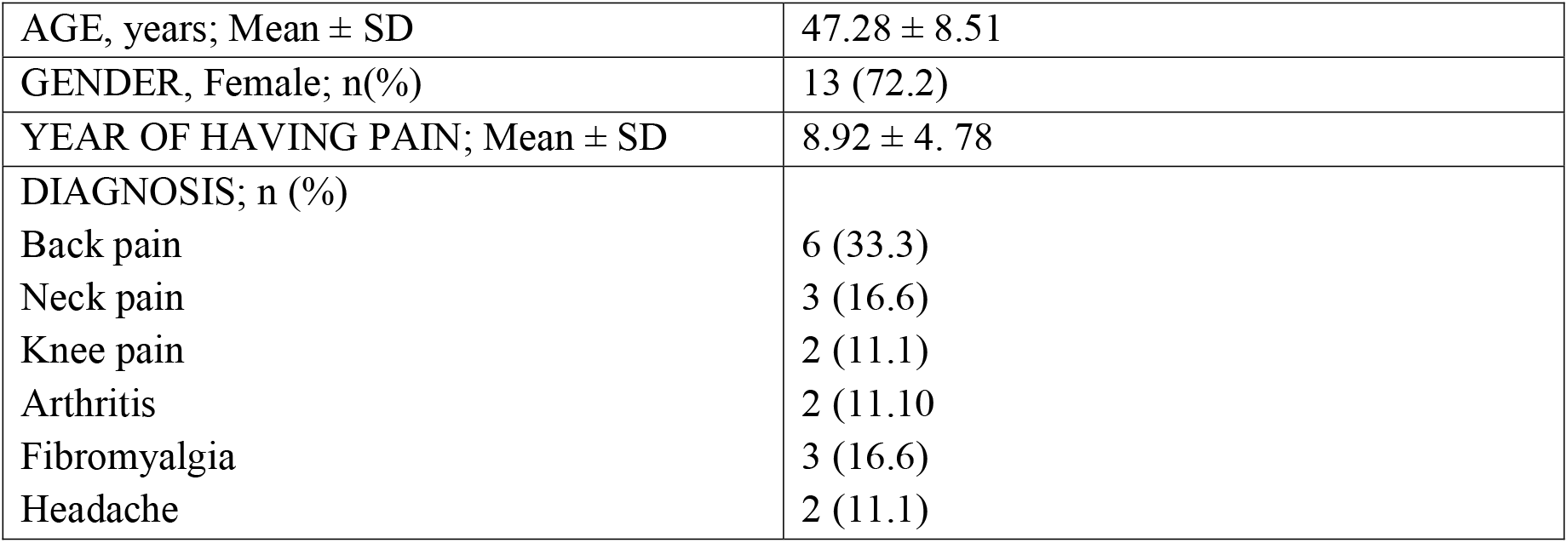
DEMOGRAPHIC CHARACTERSTICS.

|  |  |
| --- | --- |
| AGE, years; Mean ± SD | 47.28 ± 8.51 |
| GENDER, Female; n(%) | 13 (72.2) |
| YEAR OF HAVING PAIN; Mean ± SD | 8.92 ± 4. 78 |
| DIAGNOSIS; n (%) |  |
| Back pain | 6 (33.3) |
| Neck pain | 3 (16.6) |
| Knee pain | 2 (11.1) |
| Arthritis | 2 (11.10) |
| Fibromyalgia | 3 (16.6) |
| Headache | 2 (11.1) |

**TABLE 2:**
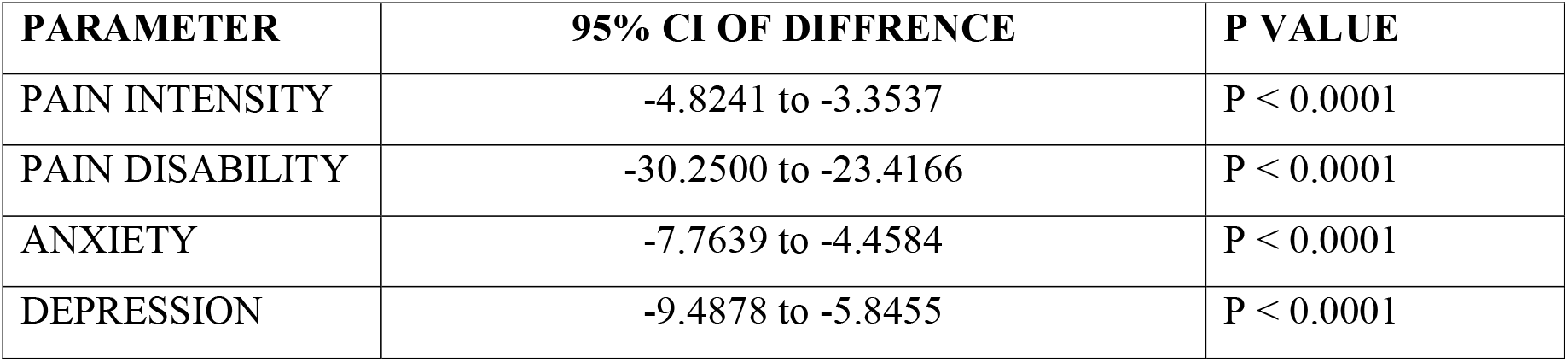
CHANGE OVER 6-WEEK OF INTERVENTION.

| PARAMETER | 95% CI OF DIFFERENCE | P VALUE |
| --- | --- | --- |
| PAIN INTENSITY | -4.8241 to -3.3537 | P < 0.0001 |
| PAIN DISABILITY | -30.2500 to -23.4166 | P < 0.0001 |
| ANXIETY | -7.7639 to -4.4584 | P < 0.0001 |
| DEPRESSION | -9.4878 to -5.8455 | P < 0.0001 |

**Figure 1:**
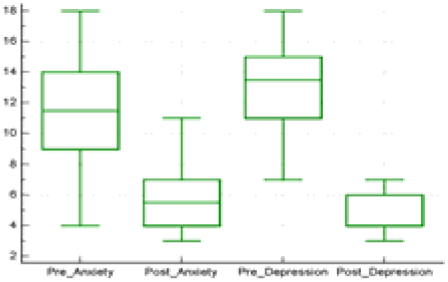
Comparison of pre and post anxiety and depression score

**Figure 2:**
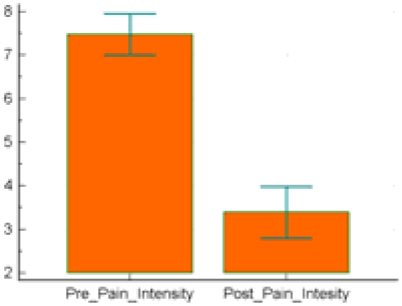
Change in pain intensity from baseline to post 6-weeks of intervention

**Figure 3:**
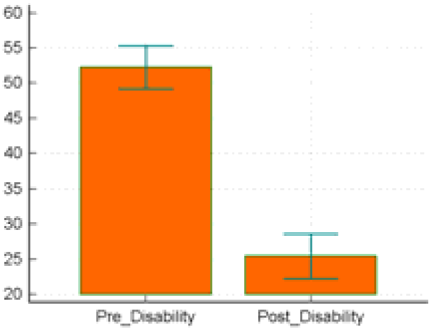
Change in Pain disability from baseline to post 6-weeks of intervention

*Paired students’ t-test was* conducted to assess the differences in means among the VAS baseline (7.48 ± 0.95), VAS post 6-week intervention (3.39 ± 1.42). The test was significant p < .0001. Also, the difference among mean values of pain disability at baseline (52.28 ± 6.14) and 6-weeks post intervention (25.44 ± 6.44) was significant p < .0001. Table 2, Graph 2 and 3 shows the change in the Pain Intensity (VAS) and Pain Disability (PDI) at baseline and 6 weeks post yoga therapy respectively.

## Discussion

Chronic pain is complex condition to treat and involves widespread issues in managing especially during pandemic. Our research demonstrates that tele-yoga therapy may reduces pain, disability and psychological symptoms associate with chronic pain.

All eighteen participants completed assessments, adhered to scheduled sessions, and completed home practices assigned. Average daily practice of 28.6 minutes every day for 4.6 days a week. Average sessions attended with yoga therapist was 11.2 sessions out of 12 over the 6-weeks. Therapists rated participants as motivated to self-manage pain, showed good understanding of the yoga principles. To our knowledge, this study is the first to deliver a tele-yoga intervention targeting chronic pain condition.

Home based intervention and the flexibility of time was valued by participants. Change in the pain intensity was observed by 2^nd^ week (by 4^th^ session) in most participants (83.3%) and that was the main motivation for continuing their own. Sixteen participants (88.8%) found self-management feasible and started to make appropriate changes on their own as pain condition changed during the week. However, problems with the technology (call interference in WhatsApp and login issues with zoom) were major challenges reported. Overall, technological issues did not affect the intervention and practice sessions. Therapist shared the session outline and video reference to practice at home after each session.

Although our trial was open to chronic pain, most of the participants who enrolled in our study had musculoskeletal pain disorder. Our findings demonstrate that we were able to recruit, screen, and deliver treatment through telecommunications, including those who had significant impairments in their daily activity participation and psychiatric functioning.

In our small sample, we found significant and sustained improvements in primary outcomes of pain intensity and impairment dur to pain. Participants also reported satisfaction with the therapy program.

Technology-delivered yoga interventions have previously demonstrated acceptable and appropriate intervention for heart failure and COPD^17^. Technology based primary care management^18^ and psychological intervention (CBT) has been found effective for chronic pain^19^, we believe that yoga also had been successfully delivered via technology, and this is an important direction for future research. Small sample size and no control group are major limitations of the study. Further large clinical trial is needed to confirm the findings.

## Conclusions

These preliminary data indicate that tele-yoga therapy was effective with chronic pain and associated disability, anxiety, depression. Future randomized controlled trials are needed to test tele-yoga treatment efficacy on chronic pain measure.

## Data Availability

The datasets generated during and/or analysed during the current study are available from the corresponding author on reasonable request.

## Acknowledgements

Authors thank all participants who shared their experience in the study. We also thank community organisation, Hindu Swayamsevak Sangh (UK) and Leicester Ageing Together for their support and participation in executing the study.

